# “I don’t know what to do or where to go”. Experiences of accessing healthcare support from the perspectives of people living with Long Covid and healthcare professionals: A qualitative study in Bradford, UK

**DOI:** 10.1101/2022.08.04.22278204

**Authors:** Sarah Akhtar Baz, Chao Fang, JD Carpentieri, Laura Sheard

**Affiliations:** Department of Health Sciences, University of York; UCL Institute of Education, University College London

**Keywords:** Long Covid, access, healthcare support, qualitative, longitudinal

## Abstract

**Background:** In July 2022 it was estimated 2 million people in the UK have self-reported Long Covid (LC).^1^ Many people have reported not receiving adequate healthcare support. There is a lack of research which provides an in-depth exploration of the barriers faced by people with LC in accessing healthcare support. It is important to understand these barriers to provide better support, care and advice for those experiencing LC.

**Objective:** To understand the barriers faced in accessing primary, secondary and specialist healthcare support for people with LC.

**Design and participation:** 40 interviews were conducted with people with LC alongside 12 interviews with healthcare professionals (HCPs) providing LC support in Bradford, as part of a UK wide qualitative longitudinal study.

**Results:** People living with LC had a large degree of difficulty in accessing healthcare services for LC support. We categorised the healthcare access experiences of participants into five main types: 1) being unable to access primary care 2) accessing primary care but receiving (perceived) inadequate support 3) extreme persistence 4) alternatives to mainstream healthcare 5) positive experiences. There was a severe lack of access to specialist LC services. Ethnic minority participants faced a further barrier of mistrust and fear of services deterring them from accessing support. HCPs discussed systemic barriers to delivering services. Experiences were embedded in macro structural issues further exacerbated by the pandemic.

**Conclusion:** To better support people with LC the barriers faced in accessing healthcare support must be addressed. Of significance, improvements to GP access are required; especially as GPs are the first line of support for people living with LC.

**Patient and public involvement:** A PPI group is engaged at regular intervals in the project.

Box 1
Patient and Public Contribution
<u>Designing an interview schedule for people with LC</u>: The wider CONVALSCENCE research project has a PPI group involved in various work packages. The PPI group is hosted by researchers at University of West of England who have expertise in patient and public involvement. Members of the PPI group all have or had LC. After an extensive literature review^4^, a draft of the interview schedule was presented to the group via a workshop. It was then revised after incorporating their feedback and piloted by the research team for further refinement.
<u>Data interpretation workshop</u>: Following advice of the UWE researchers, the PPI group were presented ahead of time with 4 interview transcripts from the dataset and provided their interpretation of the interviews via a workshop. The theme of barriers to accessing healthcare was also highlighted by attendees, for example they discussed patients being disbelieved (particularly young people) and fragmented services as some points of interest within the transcripts.

## 1. Introduction

LC is a rapidly emerging medical condition first drawing headline nationally and internationally in 2020. In the early stages of the pandemic many medical professionals and patients reported being neglected or disbelieved about their persisting COVID-19 symptoms.^2,3^ Thus, they mobilised online via social media to create awareness of their condition. As such LC is believed to be the first illness constructed by patients.^2,4^ Despite the increasing prevalence of LC its definitions remain vague and are continuously evolving. Adopting the WHO definition, NICE states that the term LC “is commonly used to describe signs and symptoms that continue to develop after acute COVID-19. It includes both ongoing symptomatic COVID-19 (from 4 to 12 weeks) and post-COVID-19 syndrome (12 weeks or more)”.^5(p5)^ Post-COVID-19 syndrome is described as presenting with a cluster of often overlapping symptoms which fluctuate, change over time, affect any system in the body^5^ and impact “everyday functioning”.^6(p674)^ The fluctuating and persistent symptoms of LC described by people include breathlessness, fatigue, cough, fever, neurological symptoms (such as loss of taste and smell and brain fog), skin rash and chest pain.^7^ There has been an increasing emergence of academic studies exploring LC and the medical and social impacts it has on people’s lives.^7–11^

The UK has universal healthcare provision which is free for most at point of delivery.^12^ However, barriers to access are impacted by a healthcare system which has faced years of austerity, budget caps, increasing waiting times, pressurised services, backlogs, and workface shortages.^12,18^ This has been further exacerbated by the pandemic, consequently impacting people’s ability to access healthcare. COVID-19 has been said to have created a ‘perfect storm’ “interacting with and exacerbated by social, economic and health inequalities”.^12(p3)^ The pandemic has further intensified health inequalities, existing chronic health and social conditions.^12^ Healthcare services are fragmented with patients transitioning between multiple care pathways; often patients consult with GPs who act as gatekeepers to other specialist services.^13,14^ Given the complexities and uncertainties surrounding the diagnosis, treatments, and impacts of LC, it is expected that it may become a burden upon the healthcare system.^15^ Although some studies and commentary pieces have touched upon patients not being believed by healthcare professionals leading to them managing symptoms alone^7,8^, and the importance of relationship-based care^16^, little is currently known about people’s experiences of not being able to access adequate healthcare support. Rather, a few studies have touched upon the healthcare access experiences of healthcare professionals with LC.^7,17^

Existing studies do not embed LC patients’ experiences within the wider structural impact the pandemic has had on the NHS, health inequalities and consequently how this shapes access to healthcare services. This paper will present findings from the first stage of the Bradford component of a national qualitative longitudinal study. The aim of the study is to understand people’s experiences of LC, the challenges they face and how their illness develops over time. This paper aims to understand healthcare access for people living with LC, after this issue arose as a striking finding.

## 2. Methods

### 2.1 Study design and setting

This paper is based on a wider qualitative longitudinal study, which has two components. Firstly 80 self-identified non-hospitalised people with LC are being interviewed over three timepoints between 2021-23 (see Figure 1). Participants are drawn from the Born in Bradford (BiB) cohort, 5 national cohort studies hosted by UCL* and further sampling via community connections in Bradford. The BiB cohort tracks the health and wellbeing of over 13,500 children, and their parents overtime. Participants drawn from the 5 national cohort studies are geographically dispersed across the UK. The second component is 3 interviews overtime with 12-15 healthcare professionals (HCPs) and those working in/with public health supporting people with LC in Bradford. In-depth semi-structured interviews have allowed for people to share the lived experiences and challenges of having LC and for HCPs to share reflections on delivering care.

**Figure 1.**
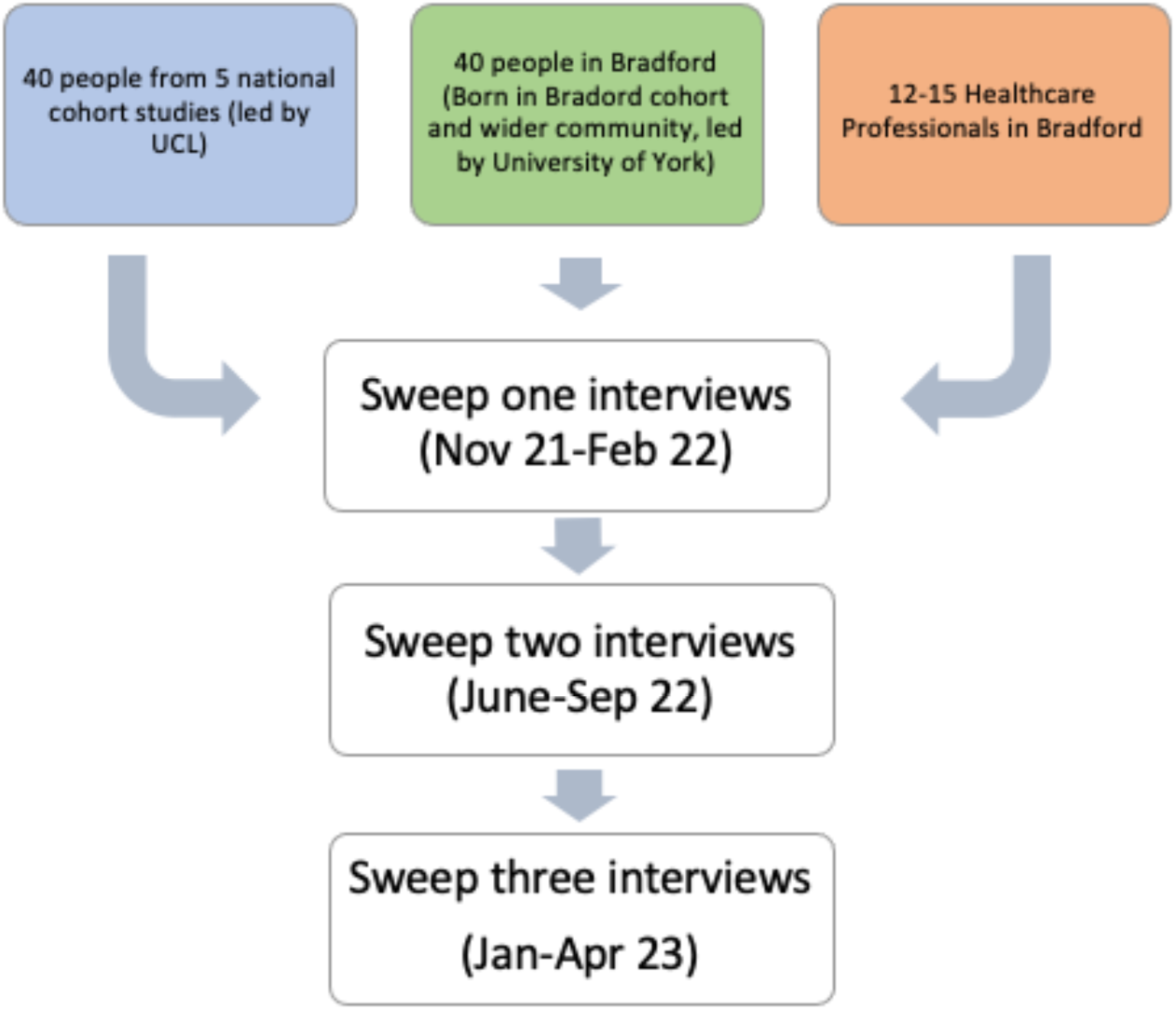
Qualitative longitudinal project timeframe.

This paper will focus on the first set of interviews of the Bradford component of the study. Bradford is a city in the North of England with high levels of deprivation, poverty and health inequalities.^19^ As such, we engaged with a socially and ethnically diverse sample. Bradford experienced a high number of COVID-19 cases compared to the rest of the UK. This is cited as “likely to be due to greater deprivation, high population density and a higher-than-average number of multi-generational households”.^20(p1160)^ Furthermore, it has been found in a racial disparities report that ethnic minorities have been overexposed to and under protected against COVID-19.^21,22^ People from deprived localities are also more vulnerable to COVID-19 infections, both groups could disproportionately experience LC.^8,12^

### 2.2 Sample and data collection

#### People with LC

Interviews were conducted with 40 people living with LC in Bradford. Sampling purposively, we aimed to oversample ethnic minorities and those living in medium to high deprivation, using postcode and IMD score as a proxy for deprivation status. We approached people with a range of engagement with healthcare services and considered the severity of LC (mild to severe self-defined symptoms). BiB cohort participants were largely in their 30s and 40s. Our sister study at UCL covers a diverse range of age groups therefore age was not a key sampling criterion here. 21 participants were drawn from the BiB cohort identified via a recurrent cohort survey. From Feb-Aug 2021 survey respondents have been asked if they have had COVID-19 and how long their symptoms lasted, with four options to choose from. Reflecting the literature at the time, although there was no firm definition of LC, it was understood to be defined as having persistent symptoms for over 4 weeks.^9^ We therefore sampled respondents who stated that their symptoms were either 5 to 12 weeks or over 12 weeks in duration. Once the list of potential participants was generated by BiB, a research assistant called respondents inviting them to take part. Information sheets and consent forms were sent out to those interested. The first author arranged the interviews. A remaining 19 people were recruited outside the cohort through community workers and snowball sampling. Those embedded in community settings had established trust and rapport with local people which allowed for a diverse range of respondents to be approached.^20^ Snowball sampling was used to take a more targeted recruitment approach and engage with underserved groups, for example when recruiting people whose first language is not English and recruiting more men.

Participants were predominately female reflecting there being more mothers registered in the BiB cohort than fathers and females being cited as having a higher risk of developing LC.^23^ Participants came from 10 different postcodes dispersed across Bradford. They worked in a range of occupations, from low-paid/low-skilled jobs, such as warehouse workers, and professional occupations such as nurses. The timeframe of when participants had their initial COVID-19 infection was broad. A few participants had COVID-19 at the very start of the pandemic when testing was not available. The rest of the participants had a confirmed infection via a PCR or lateral flow test as and when testing was available. Participants were at different stages of their LC illness; some had recovered, and most were still experiencing symptoms. At the time of the interview, there were a range of durations for which participants had LC, from being having LC for 6 weeks to around 20 months. There were a wide range of persistent symptoms reported with a loss of taste and smell, fatigue and breathlessness being common

The interviews took place between November 2021 and March 2022. All interviews were conducted remotely by the first author either over the phone or video call. Interviews in Urdu or Mirpuri were conducted with 3 people. The interviews ranged from a duration of 16 minutes to almost 2 hours (average length of 38 minutes). All participants gave informed consent via verbal audio recording at the start of interviews. Interviews were recorded digitally and transcribed by a professional transcriber with identifiable information removed. Interviews in Urdu or Mirpuri were transcribed by the first author, with data used in outputs translated over to English.

#### Healthcare professionals

HCPs were recruited starting off by contacting existing HCP contacts of the last author, followed by further snowball sampling and identification of HCPs via already recruited participants. Emails/letters were sent with an information sheet and details of what was involved. A range of HCPs and people working with/in public health running and supporting LC services in Bradford, a key criterion for recruitment, were identified. 12 decided to take part spanning from a range of primary care and LC specialist services (see Table 2). Remote interviews via video or phone call took place from December 2021 to April 2022. Again, these were recorded digitally and transcribed professionally. All interview IDs in this paper are not known to anyone outside the research team.

**Table 1:**
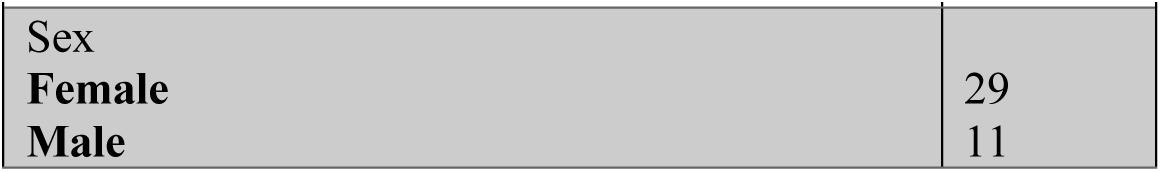
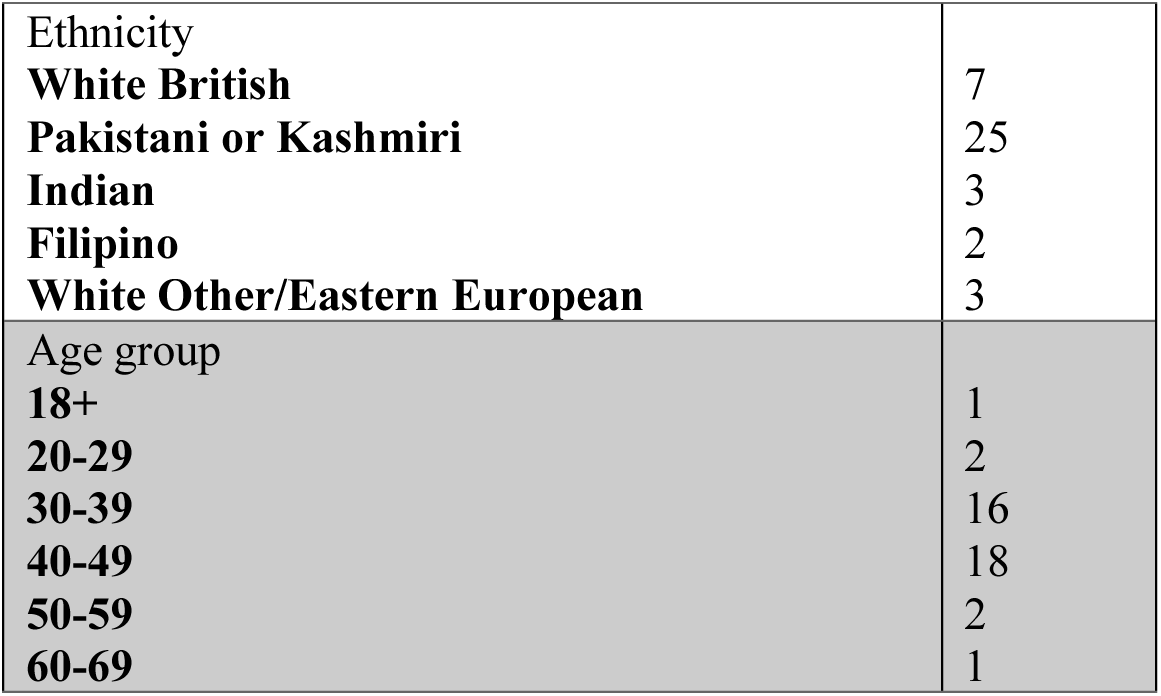
Participant demographics.

| **Table 1: Participant demographics** |  |
| --- | --- |
| Sex |  |
| Female | 29 |
| Male | 11 |
| Ethnicity |  |
| **White British** | 7 |
| **Pakistani or Kashmiri** | 25 |
| **Indian** | 3 |
| **Filipino** | 2 |
| **White Other/Eastern European** | 3 |
| Age group |  |
| **18+** | 1 |
| **20-29** | 2 |
| **30-39** | 16 |
| **40-49** | 18 |
| **50-59** | 2 |
| **60-69** | 1 |

**Table 2:** Profile of HCP participants.

| **Table 2: Profile of HCP participants** |
| --- |
| 1. Physiotherapist |
| 2. Medical practitioner working in a LC clinic |
| 3. Medical practitioner working in a LC clinic |
| 4. Occupational therapist working in a LC clinic |
| 5. GP |
| 6. Physiotherapist working in a LC clinic |
| 7. Public health official |
| 8. Charity CEO involved in health and social care |
| 9. General manager |
| 10. Senior psychologist working in a LC clinic |
| 11. Project manager |
| 12. GP in a deprived locality |

### 2.3 Analysis

A reflexive thematic analysis approach was taken.^24^ Consistent analysis sessions were held by the research team (all authors) to develop themes. Healthcare access arose as a striking finding during our initial analysis sessions, with most participants discussing substantive, lengthy content about their experiences of accessing - or failing to access - various elements of the healthcare system. This participant driven content about healthcare access continued to dominate interviews as fieldwork proceeded. We developed a coding framework which focused on healthcare access. The first author then coded the Bradford transcripts, sense checking with the other authors. The first author conducted further reflexive and interpretative work to write up the paper and analysed HCPs perspectives in relation to the access theme. The interviews were analysed in Microsoft Word without any software package.

## 3. Findings

We were immediately struck by the difficulty the majority of participants faced when accessing healthcare support for LC. This was a vociferous and very clear overarching narrative which proceeded throughout data collection. We applied the following research question to the data in order to help us make sense of what participants were telling us: “What happens when patients try to access care and support for their LC symptoms?” We found this could be delineated into three themes. First, the differing experiences that participants had of healthcare access, which we break down into five main types. Second, experiences of mistrust and fear amongst ethnic minorities in our sample. Third, systematic barriers to service delivery which was an issue discussed predominantly within the HCP interviews.

### 3.1 Experiences of accessing healthcare

We found five main types of experience which participants discussed when accessing or trying to access healthcare support for LC. First, some people with LC were not able to get through to primary care and not able to secure a GP appointment. Second, many were able to access primary care but did not receive (perceived) adequate support from either their GP or secondary care. Third, a small group of participants who were extremely persistent in their interaction with healthcare to seek LC support. Fourth, a group who used alternatives to accessing mainstream healthcare for various reasons. Fifth, a small number of people who had positive experiences. In Figure 2, we have visually represented an approximate weighting for each type. We also discovered a severe lack of access to specialist LC clinics.

**Figure 2.**
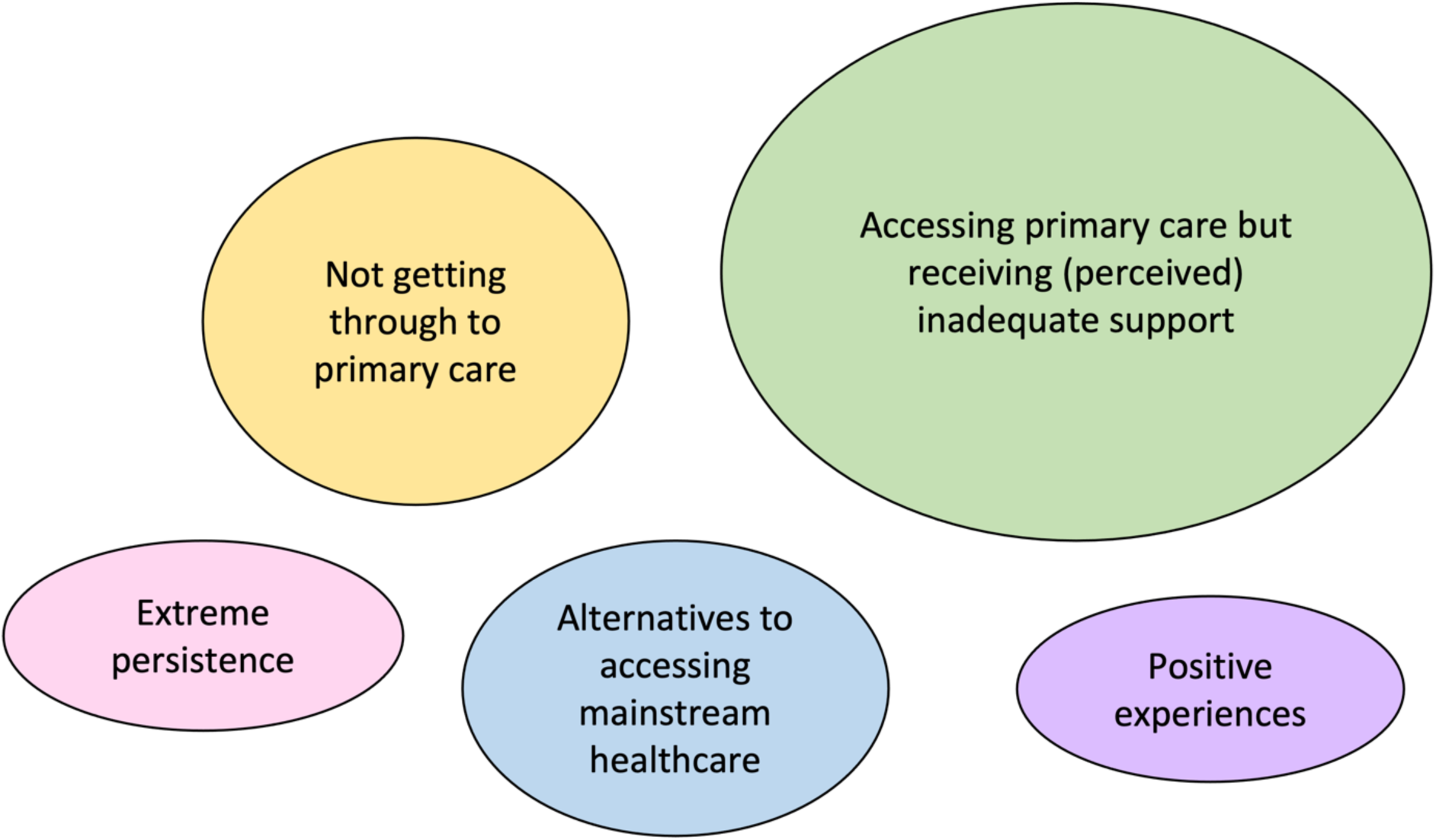
Weighting of groups (5 different experiences of healthcare)

#### Not getting through to primary care

Most notably some participants were falling at the first hurdle when trying to access support and advice for LC symptoms from GP practices, often a first point of contact for patients. A common barrier was not being able to get through to practices via the phone, often facing a prolonged wait for someone to pick up, as this extract illustrates:

> *“I will ring them and then I’m waiting on my break for like 10 minutes and nobody is answering, so I’ll wait another 20 minutes. When I’m at home and I’ve got a day off, I don’t know where to start. So I don’t want to ring my doctor waiting, you know, 2 hours on the phone because I’ve got no time for it and I’m trying to manage my symptoms with ginger or garlic*.*“ (Interviewee 11, early 40s, female, Eastern European)*

One participant was already aware that her GP was *“extraordinarily difficult”* to get through to and instead went to her local pharmacist for advice:

> *“I spoke to the chemist because our GP is extraordinarily difficult to get through and it’s very difficult to talk to anybody other than the receptionist. So I thought I’d just go and talk to our local pharmacist and see if they can suggest anything and they just said that I’ve just got to let the symptoms come out naturally or take paracetamol for my headaches*… *relax. there was nothing else offered if they could offer anything else I don’t know “ (Interviewee 29, late 30s, female, White British)*

HCP interviewees also acknowledged prolonged waiting time to get through to services as a key barrier. Resultingly, people could end up self-managing potentially risking further health complications:

> *“I think there are going to be a lot of people who we’re not touching. I mean it’s how do you get hold of your GP? Last time it took 50 phone calls, 50 tries on my mobile. How do you do that if you’re exhausted?” (HCP1, physiotherapist)*

A further barrier for this group was having to justify their need for an appointment with the receptionist, often facing push back. For example, when interviewee 34, a British Pakistani male in his early 30s, contacted his GP he felt that he was not a priority. He mentioned the LC clinic to the receptionist however had to face a long waiting time of 3 weeks for an initial appointment with the GP before referral, leaving him to state: *“So in my mind at that time it was just kind of that natural response to when you’re being pushed back to say, ‘okay I’ll leave it then’ and that was that*.*“* Thus, being able to get a GPs appointment in the first instance is one major barrier many people with LC are facing. But those who were able to eventually get through also faced hurdles within the healthcare system.

#### Accessing primary care but receiving (perceived) inadequate support

Most participants were able to get through to their GP and received an appointment but felt they had received (perceived) inadequate support from primary care. An interview with a couple with LC, living in a deprived area of Bradford, provides one account of such experiences. The difficulty of being able to access their local GP was further exacerbated during the pandemic. When they finally got through, they were not happy with the advice given:

> *Wife: “ They said just take paracetamol*.*“ Husband: “This is normal. This is common in here. Take paracetamol and ring two months*.*“ (Couple interview, 40s, Pakistani)*

Some participants described a sense of disappointment in primary care. One participant stated that he felt *“hopeless”* and *“neglected” (Pakistani male, mid 40s)*. Participants wanted to receive more advice and support from their GPs. Another participant felt that LC was not taken seriously compared to other medical conditions, a common finding reported in previous studies:^8,25^

> *“. when I spoke to the doctor’s about feeling rubbish “oh well it will be just Long Covid but we don’t do anything about it”. If I’d said “oh it’s anaemia”, then they do all these tests and you can progress. But if it’s Long Covid it’s just “well that’s what [it] is*.*” (Interviewee 20, early 40s, female, White British)*

Two participants stated that they had been referred to secondary care services by their GP but were still on a waiting list after many months. Interviewee 38 had been referred to E.N.T. and was on the neurology service waiting list for 6 months but still had no answer regarding why she was experiencing persistent head and ear pains for over a year. From the interview there was a clear sense of frustration about how long it was taking to get an appointment and navigating a fragmented healthcare service. She wanted to find answers about the cause of symptoms experienced since her COVID-19 inflection and had not been diagnosed with LC. The participant contemplated a private healthcare check-up as an alternative when visiting India to see family. There she felt she would be able to get all the tests needed in one hospital visit and find some answers:

> *“GP is also waiting for investigations and they’re just giving the medications but at the end of the day I mean I’m anxious, I don’t know what’s happening*… *Wherever I go, whatever they do they’re saying everything is fine. My chest x-ray is fine*… *So nobody knows. They’ve not diagnosed it*… *I don’t know what to do. Maybe I will go*… *back home in India*… *if I get a chance when I go back that maybe I’ll go for proper treatment So think how many months I’m just waiting for this, you know, they could have done that CT head[scan] when I was actually there. They said no, E*.*N*.*T is only doing certain parts I don’t know what to do or where to go, to whom to ask and nothing is easy access. It takes forever* …*I’m really fed up with this it’s really hopeless. I’m trying to live with it now*.*“ (Interviewee 38, early 40s, female, Indian)*

#### Extreme persistence

We found that a high level of persistence and familiarity with approaches to get through to the right person were required to gain access to primary care. A few participants were persistent in navigating their way through services to gain medical support. These were those working in professional occupations, for example public health, or those who had extensive previous experiences of navigating GP services because of other long-term illnesses. Thus, they had high health literacy and access to resources. They too acknowledged the difficulties of getting through to GPs over the phone and illustrated the importance of making sure that people get through to a GP who knows their medical history and that they access continued care overtime from the same practitioner^16^, as this extract shows:

> *“First of all they put you on a triage list and get someone to call you back and I’ve had to insist and say, I need to be put on my doctor’s list for her to ring me. There’s no point in anybody else ringing me because they don’t know me. I think that’s the bugbear isn’t it sometimes I’ve had to speak to other doctors, but they’ve not really known and you get mixed messaging. I just need to speak to my own because it’s having that trust in somebody as well isn’t it. But on the whole, I don’t have any quibble they’ve genuinely been supportive*.*“ (Interviewee 10, late 40s, female, White British)*

Furthermore, interviewee 20 who has rheumatoid arthritis discussed the resources that she drew upon to access primary care and be listened to:

> *“. I think I probably I’ve got a bit more access than most people because of my rheumatology team. They do listen you see and because of my medication, you know, they have to listen to me. Whereas if I hadn’t have had that communication and opened to me, I’m not sure it would have continued*.*“ (Interviewee 20, early 40s, female, White British)*

#### Positive experiences of healthcare

A few participants described having an overall positive experience of engaging with primary care for LC support. This included GPs listening, providing reassurance, practical and emotional support, receiving continuous care and follow-up phone calls:

> *“What I did appreciate was that my telephone call with the GP was probably slightly extended to the other ones that I have had in the past and the fact that it was the same person that I spoke to There’s an element of continuity of care that really helps” (Interviewee 1, mid-30s, female, Pakistani)*

A participant who was initially hospitalised for COVID-19 described receiving follow-up support from her GP, who provided practical advice on breathing exercises to help with continued experiences of breathlessness:

> *“For my breathing, I spoke to my GP and he recommended me to like get balloons and kind of blow into them. Breathing exercises. So I used to do breathing exercises*…*“ (Interviewee 5, 18+, female, Pakistani)*

Overall, such approaches were cited as being helpful in managing the illness and can be learnt from to provide better support to people with LC.

#### Using alternatives to accessing mainstream healthcare

Every participant described a degree of self-management or ‘burden of illness’^26^, for example prioritising rest, reducing physical activity, or using home remedies. However, some participants self-managed symptoms from the offset and chose not to engage with healthcare services. This was due to several reasons including, not preferring to approach healthcare services unless necessary, not liking medicine, preferring self-management, not wanting to burden an already overwhelmed NHS, not knowing what help was available, mistrust, fear and past negative experiences which deterred healthcare access (see section 3.2) and learning to live with symptoms with the hope that they would see change overtime. One HCP stated that some may be *“accepting that this is how life is for them” (HCP5, GP)*.

#### Limited access to LC clinics

A startling finding was that only 1 out of 40 interviewees in Bradford had engaged with a LC clinic service. This one person was an NHS staff member and had accessed an LC clinic through their workplace that was designed to help NHS staff recover and progress back to work. We interviewed the ‘average person’ in Bradford, many had not even heard of a LC clinic. A very small proportion were beginning to discuss the possibility of a referral with their GP if symptoms worsened.

Patients have to go through a prolonged process with their GP to gain referral to the LC clinic. A clinician interviewee stated that to gain access to the clinic symptoms must last for 12 weeks or more. Patients must go through an initial assessment with their GP to eliminate other health risks. It was argued that this timeframe can be reduced so people can receive earlier interventions:

> *“I think probably identifying the right people, you know, so making sure that people don’t miss out. I think probably not being necessarily so strict about this 12-week cut off, you know, because even at the moment the GPs are not allowed to refer to the community team until it’s 12 weeks. But why not refer at 7 weeks, you know? Why wait?” (HCP2, medical practitioner)*

Evidently participants had to do the ‘hard and heavy work’^8^ to receive healthcare support for their symptoms. This depicts the different barriers and inequalities in accessing services amongst participants. The next section will further focus on mistrust and fear as an extra layer of barrier to accessing services for ethnic minorities.

### 3.2 Mistrust and fear

Mistrust and fear were a pertinent issue amongst some ethnic minority participants. This has previously been cited as a barrier in relation to COVID-19 vaccine uptake amongst ethnic minorities^20^ and creates an additional bottleneck for people with LC. A few participants expressed fear of going to the hospital for treatment of their LC symptoms. In one participant’s case, this reflected ‘fake news’ stories and rumours going around the Pakistani community in Bradford at the height of the pandemic that hospitalisation could lead to death.^20^ Interviewee 4 emphasised a lack of trust in doctors and a need to increase trust in healthcare services to tackle such rumours. This can be embedded in both experiences of historical and contemporary structural racism which leads to mistrust in the healthcare system and may be further exacerbated in the case of COVID-19 as there has been a disproportionate number of deaths amongst ethnic minority people.^27^

> *“*…*you should have that full trust in him [GP]*…*this ‘negativity’ that is spread this this this should not happen because I understand because I I didn’t go to the hospital I didn’t go because of this that I heard that that’s it if you go to the hospital then a person does not come back alive*…*“ (translated from Urdu) (Interviewee 4, late 30s, male, Pakistani, migrant)*

The participant stated that he attained medical advice informally from a GP, which a family member put him in touch with. The GP spoke his preferred language, provided reassurance, advice and *“emotional support”*.

Other participants also expressed fear of being hospitalised, put on a ventilator, and dying. This also reflects some participants knowing of people who have died from COVID-19 making it more ‘real’.^20^ This was deterring interviewee 36 from seeking medical support when experiencing frightening pains in his chest due to his LC. He was yet to take the first step to engage with services:

> *“I felt so wheezy and I felt like my chest was tightening up around me and I was really close a few times to making the call to 911[111] to say, you know, this is happening, what should I do and I couldn’t do it because I was too scared to make the call*.*“ Interviewer: “In what ways were you scared?” “I think like I try not to listen to people but I heard a lot of stories at the time that people were going to the hospital and not coming back out and were put on ventilators and stuff. That essentially was really scaring me and I did have one of my close friends, his brother passed away*… *Now that didn’t scare me but it kind of puts that thing in your mind. He wasn’t vaccinated. I’m vaccinated. But yeah just things like that really. I don’t want to put myself in that position*.*“ (Interviewee 36, mid-30s, male, Pakistani)*

There were also accounts of participants lacking confidence and trust in HCPs. This was embedded in previous encounters with GP surgeries where they were misbelieved or not taken seriously. These past encounters played a part in deterring them from seeking medical advice for LC. These experiences occurred at the intersection of aged, gendered and racialised discrimination. For example, a young Pakistani woman described not being taken seriously by her GP:

> *“I mean my doctors aren’t really that good in that sense anyway, so I wouldn’t even go to them for help*… *I went to the doctors once because I had a lump on my breast and he told me to lose weight. They didn’t even check. In that kind of sense I don’t go to the GP anymore because of them not being really practical about anything*… *it stops me*…*“ (Interviewee 30, 20s, female, Pakistani)*

Another Pakistani man in his early 30s described feeling that as a *“youngish man”* he was not prioritised or taken seriously and was previously *“denied”* being given antibiotics for a medical condition, with his practice stating: *“you’re a young fit guy, you’ll be fine”*. Therefore, mistrust ran in both directions as patients have been mistrusted by HCPs which consequently shaped their mistrust of the system.

Often in relation to ethnic minority experiences of healthcare access language is cited as the key barrier. However, the 3 Urdu/Mirpuri speaking participants in this study stated that their family, husband, or support networks supported them in seeking medical advice. This raises the importance of shifting the conversation beyond a narrow focus on solely language to other barriers, namely mistrust and fear. This creates an additional barrier to access for ethnic minority people despite language abilities. Past encounters of being disbelieved, having mistrust and fear can lead to a lack of confidence that adequate healthcare support will be provided, consequently impacting people’s decisions on seeking support for LC symptoms, resulting in people self-managing symptoms.

### 3.3 Systemic barriers to service delivery: HCPs perspectives

HCPs in Bradford shared their perceptions about the barriers people with LC face when accessing healthcare. There was a mix of both praise and criticism of services. However, a salient finding was the systemic healthcare access issues that HCPs had to work around. Firstly, there was a lack of training for GPs about LC, particularly during the onset of the illness. HCPs were overstretched and often had to figure out themselves what LC was and how to support patients, drawing on knowledge of other illness like chronic fatigue syndrome, and in one GPs case via her own experience of LC:

> *“*…*there was nothing to offer so we were kind of winging it*… *making sure we weren’t missing our you know bread and butter stuff erm but it just kind of felt like there was something happening to these patients that we didn’t know what was happening*… *it was something I was reading a lot about*…*“ (HCP12, GP working in a deprived locality)*

As discussed in section 3.1., there was limited access to specialist LC services, with an emphasis being placed on access for NHS staff with LC and patients who had been hospitalised. A physiotherapist was informed to use existing services in her own practice to support people with LC, despite already being overstretched and with increasing workloads. Nevertheless, she continued to support her LC patients:

> *“we do more than we should and erm we work more and more and later and later and then we cannot fill all our workload*… *we all know that in reality it means services are overstretched ones that are already overstretched*…*“ (HCP1, physiotherapist)*

At the time of fieldwork, a newly set up LC clinic aimed to provide holistic care to Bradford patients taking a multidisciplinary approach, which is particularly key as LC is often seen as a primarily respiratory phenomenon. The main barrier to accessing this clinic was the long waiting list. One HCP stated that the clinic was *“lagging behind”* due to the time it had taken to set-up and allocate funding. Additionally, there has been a struggle to recruit staff due to shortages of specialist staff, a wider system issue which is also impacting this service.^18^

Although HCPs felt that GPs were best equipped to support LC patients, as they had knowledge of the whole body, a CEO of a third sector organisation working with health services raised the concern of accessing the clinic via GPs, the primary route of referral. This can create an immediate *“bottleneck”* as many face barriers to accessing GP appointments, particularly marginalised groups. This further illustrates the high importance of improving access to primary care but also to use other methods to signpost patients to specialist services, including more engagement in grassroot community settings that are connected to the most underserved:

> *“if there was a way for a wide range of groups to be able to refer, connect, signpost people to that service without having to jump through hoops for a GP then I think that will be more effective” (HCP8, charity CEO)*

One GP working in a deprived locality stated that some of her patients were now able to access the clinic and shared positive experiences. However, she was yet to receive any information from the clinic on the progress made and instead had to ask patients. This reflects a fragmented healthcare system where patient record systems are not linked together between services, creating barriers to better supporting patients.^28^

Importantly, these findings illustrate the impact of systemic issues on service delivery and the access and support people with LC get from HCPs.

## 4. Discussion and implications

This study offers a more in-depth insight into barriers faced by people with LC in accessing healthcare support. We found five different types of experience when accessing healthcare alongside a lack of access to LC specialist services. Overall, it is evident that people faced worrying difficulties in accessing the healthcare system at all and a high degree of persistence was required just to access primary care. There were some positive experiences of primary care such as GPs following up, but many participants felt that their symptoms were not taken seriously as cited in previous studies.^28^ People who were referred to secondary care had to wait many months to access services. Only 1/40 interviewees had accessed a LC specialist service, with a few people discussing the possibilities of future referral with their GP. It is important to embed these subjective experiences in a structural context. Backlogs, a decimated and underfunded healthcare system, and workforce shortages mean people with LC experience a bottleneck in accessing services.^7,14,29^ Access to primary care is particularly difficult for those living in deprived postcodes where there is higher prevalence of inequalities and ill health, therefore more demand for services.^12^ This can be frustrating when people are managing a new illness for which there is no current holistic treatment. People with LC have been previously described as having to do the ‘hard and heavy work’ of both understanding and managing a new illness and navigating fragmented healthcare services.^8,11^ This was certainly evident in our interviewees’ accounts.

Evidently, GPs are often the first point of contact for patients and play a crucial role as gatekeepers in facilitating access to secondary care and LC clinics, and assessing patients.^14,25^ Therefore, it is essential to improve access to primary care so people with LC are provided with better support and referral. Our study shows that this is a major barrier for LC sufferers, this was emphasised by both people with LC and HCPs. Furthermore, this research adds further to emerging literature surrounding COVID-19, ethnicity, and mistrust amongst ethnic minorities.^15,20,30^ In relation to accessing LC support, experiences of mistrust and fear were rooted in the disproportionate impact of COVID-19 on ethnic minorities and previous negative encounters with the healthcare system.^31^ This created an additional layer of barrier in accessing healthcare support for LC amongst this group of participants. This must be considered and addressed when looking at access to and engagement with healthcare services, particularly as Bradford has a diverse ethnic minority population. Although progress has been made in setting up a LC clinic, HCPs cited the structural barriers in the healthcare system which impacted their ability to provide support to LC sufferers. As previously cited^13,28,32^ better communication between fragmented services is required so GPs can provide better follow-up support, alongside more training and education for HCPs about LC. Wider systemic issues routed in years of austerity are evidently also impacting access. There is concern that not everyone is able to seek help in an overwhelmed system.

## 5. Strengths, limitations and further research

Strengths: Qualitative LC studies, to date, have predominately focused on the experiences of mainly White populations drawing their sample from online LC support groups and healthcare professionals.^3,17,25^ In our study, we interviewed the ‘average-person’ with LC in Bradford. As such, our sample was 75% ethnic minority and mostly lived in deprived areas which allowed us to understand how the experience of healthcare access is shaped for this group of people. A key strength of the study is its three timepoint longitudinal element. The data reported in this paper are from the first set of interviews and in the next two timepoints, we will explore whether participants have engaged with and accessed further support from healthcare services. Another key strength of the study is that it also explores HCPs perspective of LC service delivery, addressing a significant gap in the literature.

Limitations: In this paper we only focus on the experiences of LC patients in one place, Bradford. In future analyses, we intend to combine the data from the Bradford sample and the national sample from the five cohort studies to understand varying topics of importance to participants. The data reported represent only one snapshot in time in a quickly evolving field, hence LC services on the ground could look different at our next timepoint. Lastly, we do not have ‘evidence’ of COVID-19 infection for some participants, particularly those who were infected during Spring 2020 when testing was largely unavailable. This could be viewed as a limitation but, rather, we see the self-identification of our sample as a positive move echoing Alwan’s assertion that “the burden of proof should not be on ill people”.^33(p201)^

## 6. Conclusion

This paper has contributed to providing a more nuanced and in-depth understanding of the barriers and ‘hard and heavy work’^8^ people with LC face in accessing healthcare support, drawing on the perspectives of people living with LC and HCPs. These subjective experiences are embedded within deep seated structural and systemic barriers, discrimination and health inequalities which create healthcare access barriers for people with LC.

### Note

*The 5 national cohort studies hosted by UCL consist of the Millennium Cohort Study (born 2000-02), Next Steps (born 1989-90), the 1970 British Cohort Study (born 1970), National Child Development Study (born 1958) and the National Survey of Health and Development (born 1946).

## Supporting information

ICMJE Disclosure Form PrePrint Submission - Medrxiv PrePrint SUBMISSION - I don't know what to do or where to go

## Data Availability

The data that support the findings of this study are available on request from the corresponding author. The data are not publicly available due to privacy or ethical restrictions.

## Acknowledgements

The authors would like to thank the Born in Bradford cohort, researchers at Bradford Institute for Health Research who supported the study and recruitment process (particularly Dagmar Waiblinger, Zenam Bi and Shahid Islam), the PPI group at the University of West England and Andy Gibson, John Kellas and Sophie Taysom who facilitated PPI workshops. A special thank you to all of the participants who took part in this study and shared their experiences.

## Ethical approval

This study has been ethically reviewed and given a favourable opinion by University of York Health Sciences Research Governance Committee on 01/10/21 (ref: HSRGC/2021/466/B).

## Author contributions

JD and LS designed the wider qualitative longitudinal study and obtained funding as part of the CONVALESCENCE grant. SB collected all the data discussed in this paper and was primary analyst for the healthcare access focus. All authors fed into analytic discussions. SB and LS wrote the first drafts of the paper. All authors read, commented and provided feedback on the paper, providing approval for the final manuscript.

## Appendix

